# Cluster of COVID-19 in northern France: A retrospective closed cohort study

**DOI:** 10.1101/2020.04.18.20071134

**Authors:** Arnaud Fontanet, Laura Tondeur, Yoann Madec, Rebecca Grant, Camille Besombes, Nathalie Jolly, Sandrine Fernandes Pellerin, Marie-Noëlle Ungeheuer, Isabelle Cailleau, Lucie Kuhmel, Sarah Temmam, Christèle Huon, Kuang-Yu Chen, Bernadette Crescenzo, Sandie Munier, Caroline Demeret, Ludivine Grzelak, Isabelle Staropoli, Timothée Bruel, Pierre Gallian, Simon Cauchemez, Sylvie van der Werf, Olivier Schwartz, Marc Eloit, Bruno Hoen

## Abstract

**Background:** The Oise department in France has been heavily affected by COVID-19 in early 2020.

**Methods:** Between 30 March and 4 April 2020, we conducted a retrospective closed cohort study among pupils, their parents and siblings, as well as teachers and non-teaching staff of a high-school located in Oise. Participants completed a questionnaire that covered history of fever and/or respiratory symptoms since 13 January 2020 and had blood tested for the presence of anti-SARS-CoV-2 antibodies. The infection attack rate (IAR) was defined as the proportion of participants with confirmed SARS-CoV-2 infection based on antibody detection. Blood samples from two blood donor centres collected between 23 and 27 March 2020 in the Oise department were also tested for presence of anti-SARS-CoV-2 antibodies.

**Findings:** Of the 661 participants (median age: 37 years), 171 participants had anti-SARS-CoV-2 antibodies. The overall IAR was 25.9% (95% confidence interval (CI) = 22.6-29.4), and the infection fatality rate was 0% (one-sided 97.5% CI = 0 - 2.1). Nine of the ten participants hospitalised since mid-January were in the infected group, giving a hospitalisation rate of 5.3% (95% CI = 2.4 –9.8). Anosmia and ageusia had high positive predictive values for SARS-CoV-2 infection (84.7% and 88.1%, respectively). Smokers had a lower IAR compared to non-smokers (7.2% versus 28.0%, P <0.001). The proportion of infected individuals who had no symptoms during the study period was 17.0% (95% CI = – 23.4). The proportion of donors with anti-SARS-CoV-2 antibodies in two nearby blood banks of the Oise department was 3.0% (95% CI = 1.1 - 6.4).

**Interpretation:** The relatively low IAR observed in an area where SARS-CoV-2 actively circulated weeks before confinement measures indicates that establishing herd immunity will take time, and that lifting these measures in France will be long and complex.

**Funding:** Institut Pasteur, CNRS, Université de Paris, Santé publique France, Labex IBEID (ANR-10-LABX-62-IBEID), REACTing, EU grant Recover, INCEPTION project (PIA/ANR-16-CONV-0005).

**Research in context:** *Evidence before the study:* The first COVID-19 cases in France were reported on 24 January 2020. Substantial transmission has occurred since then, with the Oise department, north of Paris, one of the heaviest affected areas in the early stages of the epidemic in France. As of 13 April 2020, 98,076 cases had been diagnosed in France, including 5,379 deaths. Epidemiological and clinical characteristics of patients with COVID-19 have been widely reported, but this has largely been centred on cases requiring medical care. What remains unclear at this stage is the extent to which SARS-CoV-2 infections may be asymptomatic or present as subclinical, non-specific symptoms. While extensive contact tracing has identified asymptomatic infections using RT-PCR testing, serologic detection of anti-SARS-CoV-2 antibodies is needed to determine the real infection attack rate and the proportion of all infections that are asymptomatic or subclinical.

*Added value of this study:* Using a combination of serologic assays with high sensitivity and specificity for anti-SARS-CoV-2 antibodies, we conducted a retrospective closed cohort study. In a high school linked to a cluster of COVID-19 in the Oise department, we showed an overall infection attack rate (IAR) of 40.9% in the high school group, and 10.9% in parents and siblings of the pupils. The proportion of infected individuals who had no symptoms during the study period was 17.0%.

*Implications of all of the available evidence:* The relatively low IAR in this area where SARS-CoV-2 actively circulated before confinement measures were introduced indicates that establishing herd immunity will take time, and that the lifting of these measures in France will be long and complex.

## Introduction

Severe acute respiratory syndrome coronavirus-2 (SARS-CoV-2) is the novel coronavirus that was first reported to the World Health Organization (WHO) as a cluster of viral pneumonia cases of unknown etiology in Wuhan, China on 31 December 2019. It is now known to cause coronavirus disease (COVID-19), which primarily affects the upper and lower respiratory tract. On 30 January 2020, WHO declared the COVID-19 outbreak to constitute a Public Health Emergency of International Concern.^1^ Since then, the outbreak has continued to spread around the world and was described by WHO as a pandemic on 11 March 2020.

The first three COVID-19 cases identified in France were reported on 24 January 2020 in travellers returning from Wuhan, China.^2^ On 24 February, a patient from the Oise department, north of Paris, was admitted to hospital in Paris in a critical condition and diagnosed with SARS-CoV-2 infection. He died on 25 February and was the first reported COVID-19 case in France without a direct or indirect epidemiological link to China. The ensuing epidemiological investigation led to the identification of a cluster of COVID-19 that involved a high school in the Oise department.

With any emerging infectious disease, including COVID-19, initial surveillance focuses primarily on severe infections, leading to overestimates of the case fatality rate (CFR). Likewise, in the absence of knowledge of subclinical or asymptomatic forms of the infection, it is not possible to estimate infection fatality rates (IFR) except through mathematical modelling.^3^ However, population-based serological investigations use serologic assays to determine infection attack rates (IAR) in the population in an epidemic area, and are able to determine the proportion of subclinical and asymptomatic infections. This information is key for a better understanding of SARS-CoV-2 circulation, more precise estimates of fatality rates, and calibration of mathematical models used to forecast the dynamics of the ongoing epidemic.

To date, access to validated serologic assays for SARS-CoV-2 is limited.^4,5^ However, a series of serologic assays recently developed by the Institut Pasteur in Paris, France have shown high sensitivity and specificity for the detection of anti-SARS-CoV-2 antibodies.^6^

Here, we describe a retrospective closed cohort study aiming at estimating the IAR and its determinants in an area affected by COVID-19, using these serologic assays.

## Methods

### Initial case and contact investigation

Following the confirmation of COVID-19 in the patient from the Oise department, case investigation and contact tracing identified two cases in a high school and who had symptoms consistent with COVID-19 on 2 February 2020, suggesting circulation of the virus in the Oise department since the end of January 2020.

### Study design

As a follow-up to the initial case investigation and contact tracing, a retrospective closed cohort study was conducted in the high school. Between 30 March and 4 April, all pupils, as well as teachers and non-teaching staff (administrative, cleaners, catering) from the high school were invited to participate in the investigation. Since most pupils were minor, at least one parent was invited to participate in the study, to provide informed consent for their child and for any of the other children over the age of 5 years in the household enrolled in the study.

Following informed consent, participants completed a questionnaire which covered sociodemographic information, underlying medical conditions, history of symptoms since 13 January 2020, and history of COVID-19 diagnosis prior to this investigation. A 5 mL blood sample was taken from all participants, irrespective of whether they had reported fever or respiratory symptoms since 13 January 2020.

### Seroprevalence investigation in nearby blood donation centres

Between 23 and 27 March, 200 serum samples were collected from two blood donation centres, located 50 and 60 km from the place of the seroepidemiological investigation.

### Laboratory analyses

All serum samples were tested for antibody responses to SARS-CoV-2 using several assays developed by Institut Pasteur : an ELISA N assay, detecting antibodies binding to the N protein; a S-Flow assay, which is a flow-cytometry based assay detecting anti-S IgG; and a LIPS assay, which is an immunoprecipitation-based assay detecting anti-N and anti-S1 IgG. Participants were considered seropositive for SARS-CoV-2 if any test was positive, since all tests had a specificity higher than 99% with the cut-offs chosen for positivity.^6^

### Case definitions

Any participant with a positive serology at the time of blood sampling was considered as a confirmed SARS-CoV-2 infection. Each infection was categorised as symptomatic if any symptoms were reported by the participant since 13 January 2020, or, alternatively, as asymptomatic. Symptoms were considered only if they occurred at least 7 days prior to the date of blood sample collection to allow time for seroconversion.^4,7^ Symptoms were further categorized as major (fever, dry cough, dyspnoea, anosmia and ageusia) or minor (sore throat, rhinitis, muscle pain, diarrhoea, headache, asthenia).

### Statistical analyses

The infection attack rate (IAR) was defined as the proportion of all participants with confirmed SARS-CoV-2 infection based on antibody detection. It was compared by age, sex, occupation, smoking, comorbid conditions and recent symptoms using chi-squared test. Logistic regression was used to adjust for age or occupation when analysing the association between smoking and SARS-CoV-2 infection.

### Ethical considerations

This study was registered with ClinicalTrials.gov (NCT04325646) and received ethical approval by the Comité de Protection des Personnes Ile de France III. Informed consent was obtained from all participants.

## Results

### Retrospective closed cohort study

From 30 March to 4 April 2020, 878 of 1262 high school pupils, teachers, and non-teaching staff were invited by e-mail to participate in the investigation (email addresses were not available for 384). Of these, 326 (37%) responded and accepted to participate in the study. An additional 345 parents and siblings of high school pupils were also invited to participate in the study. This formed a study population of 661 participants (see Figure 1). Table 1 indicates the characteristics of the 661 participants. Pupils and their parents constituted the majority of the study population (36.3% and 31.9%, respectively). The median age was 37 years (IQR: 16-47) and 251 (38.0%) were male.

**Table 1.** Characteristics of the 661 participants of the SARS-CoV-2 seroepidemiological investigation conducted in Oise, northern France from 30 March to 4 April 2020

| Characteristics | Statistics |
| --- | --- |
| Male gender | 251 (38.0) |
| Age (years), Median (IQR) | 37 (16-47) |
| Age groups |  |
| ≤14 | 37 (5.6) |
| 15-17 | 205 (31.0) |
| 18-44 | 177 (26.8) |
| 45-64 | 239 (36.2) |
| ≥65 | 2 (0.3) |
| Missing | 1 (0.1) |
| Status |  |
| Pupil | 240 (36.3) |
| Teacher | 53 (8.0) |
| School staff | 27 (4.1) |
| Parent of a pupil | 211 (31.9) |
| Sibling of a pupil | 127 (19.2) |
| Other | 3 (0.5) |
| Symptoms |  |
| None | 209 (31.6) |
| Minor only | 131 (19.8) |
| Major | 321 (48.6) |
| Detail of symptoms |  |
| Fever | 173 (26.2) |
| Cough | 234 (35.4) |
| Dyspnea | 93 (14.1) |
| Anosmia | 59 (8.9) |
| Ageusia | 59 (8.9) |
| Myalgia | 159 (24.0) |
| Sore throat | 177 (26.8) |
| Rhinitis | 253 (38.3) |
| Diarrhea | 92 (13.9) |
| Headache | 204 (30.9) |
| Asthenia | 196 (29.6) |
| Other | 127 (19.2) |

**Figure 1.**
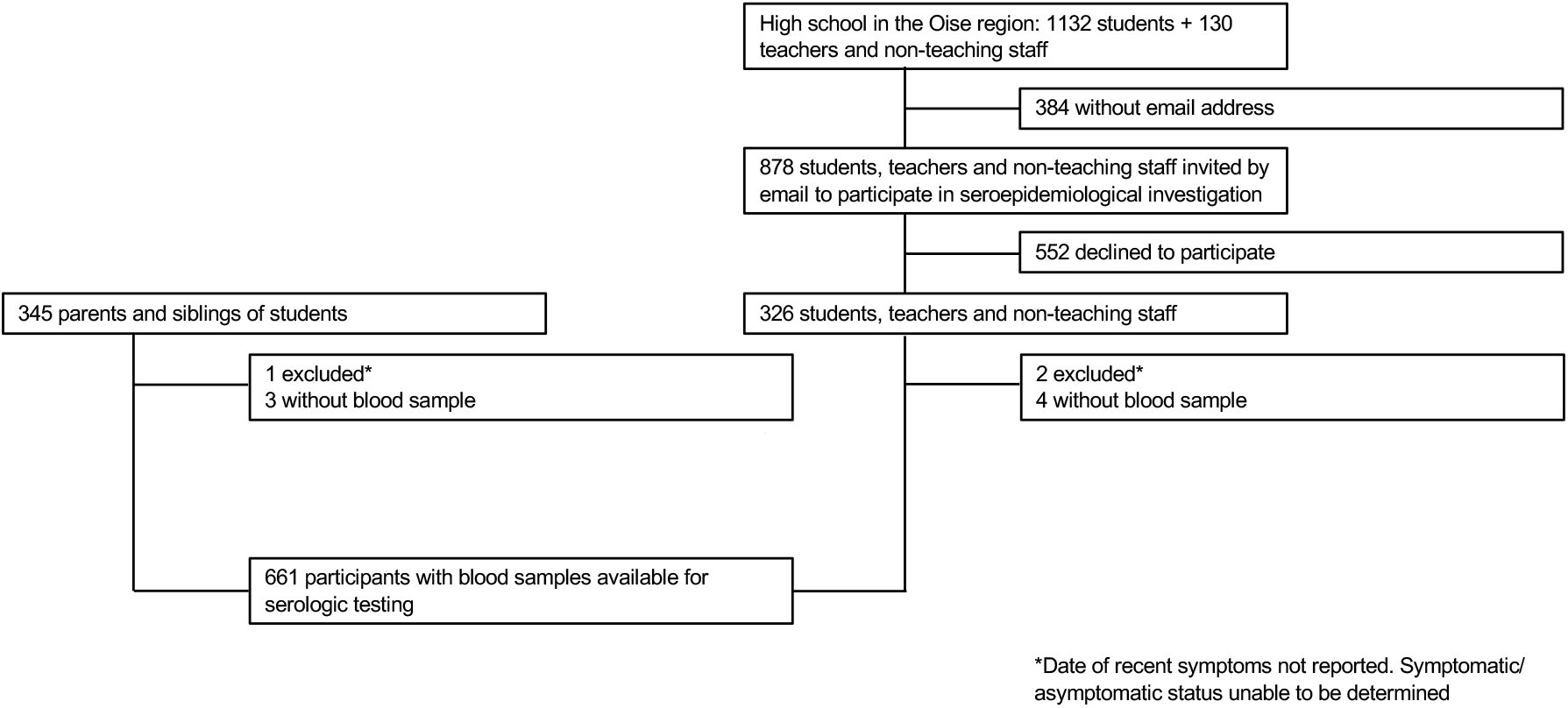
Flowchart of enrolment of participants.

Overall, 452 (68.4%) participants reported respiratory symptoms between 13 January and up to one week before blood sampling. Major symptoms had been experienced by 321(48.6%) of study participants, minor symptoms by 131 (19.8%), while 209 (31.6%) had not noticed any symptom during the period covered by the study. Most common symptoms were rhinitis (38.3%), followed by cough (35.4%), headache (30.9%), asthenia (29.6%), sore throat (26.8%), and fever (26.2%). Ten participants reported hospital admission in relation to the reported symptoms. Further investigation established no fatalities among the 1262 members of the high school population since the beginning of the study period.

Of the 661 participants, 171 participants had anti-SARS-CoV-2 antibodies (see Supplementary material), giving an overall IAR of 25.9% (95% confidence interval (CI); 22.6-29.4), and an infection fatality rate (IFR) of 0% (one-sided 97.5% CI = 0 - 2.1). Nine of the ten hospitalised were in the SARS-CoV-2 infected group, giving a hospitalisation rate of 5.3% (95% CI = 2.4 –9.8). The median age of hospitalised participants was higher compared to non-hospitalised in the SARS-CoV-2 infected group (49.0 versus 17.7 years, respectively; *P* = 0.04). Among the 171 participants with confirmed SARS-CoV-2 infection, the proportions of those with major, minor or no symptoms were 70.8% (95% CI = 63.3-77.5), 12.3% (95% CI = 7.8-18.2), and 17.0% (95% CI = 11.2 – 23.4), respectively.

Table 2 shows the proportion of those with anti-SARS-CoV-2 antibodies. There was no difference in IAR between males and females, while IAR was highest (40.0%) in the 15-17 years age group. The IAR was higher in the high school group (38.3% 43.4%, and 59.3% for pupils, teachers, and school staff, respectively) than in parents and siblings (11.4% and 10.2%, respectively) (P <0.001). Smoking was found to be associated with a lower risk of infection (7.2% versus 28.0% for smokers and non-smokers, respectively; P<0.001; OR = 0.20, 95%CI = 0.08-0.51), and this association remained significant after adjustment for age (OR = 0.23; 95% CI = 0.09 – 0.59) or occupation (OR = 0.27; 95% CI = 0.10 – 0.71). There was no increase in IAR among those who had comorbidities, otherwise known to be associated with severe forms of COVID-19.

**Table 2:**
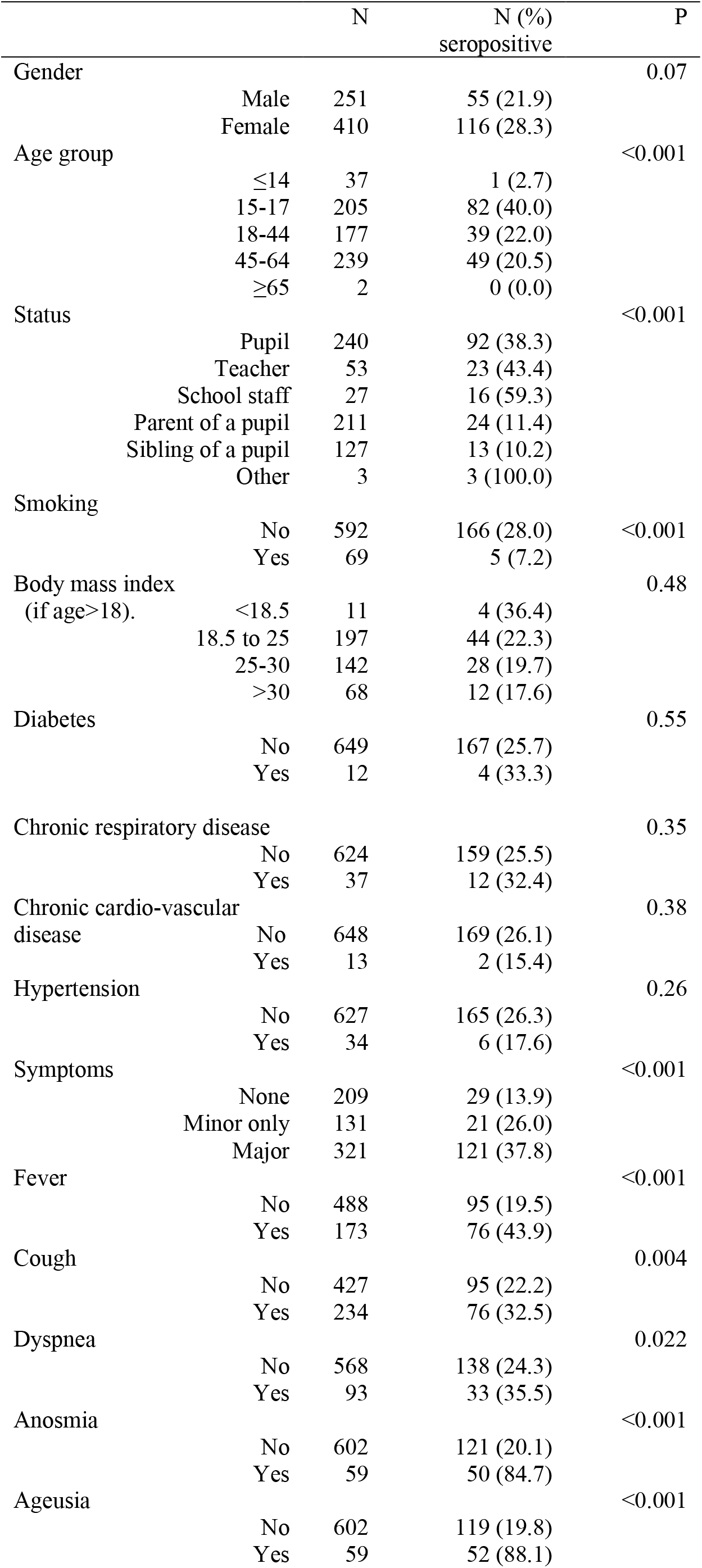
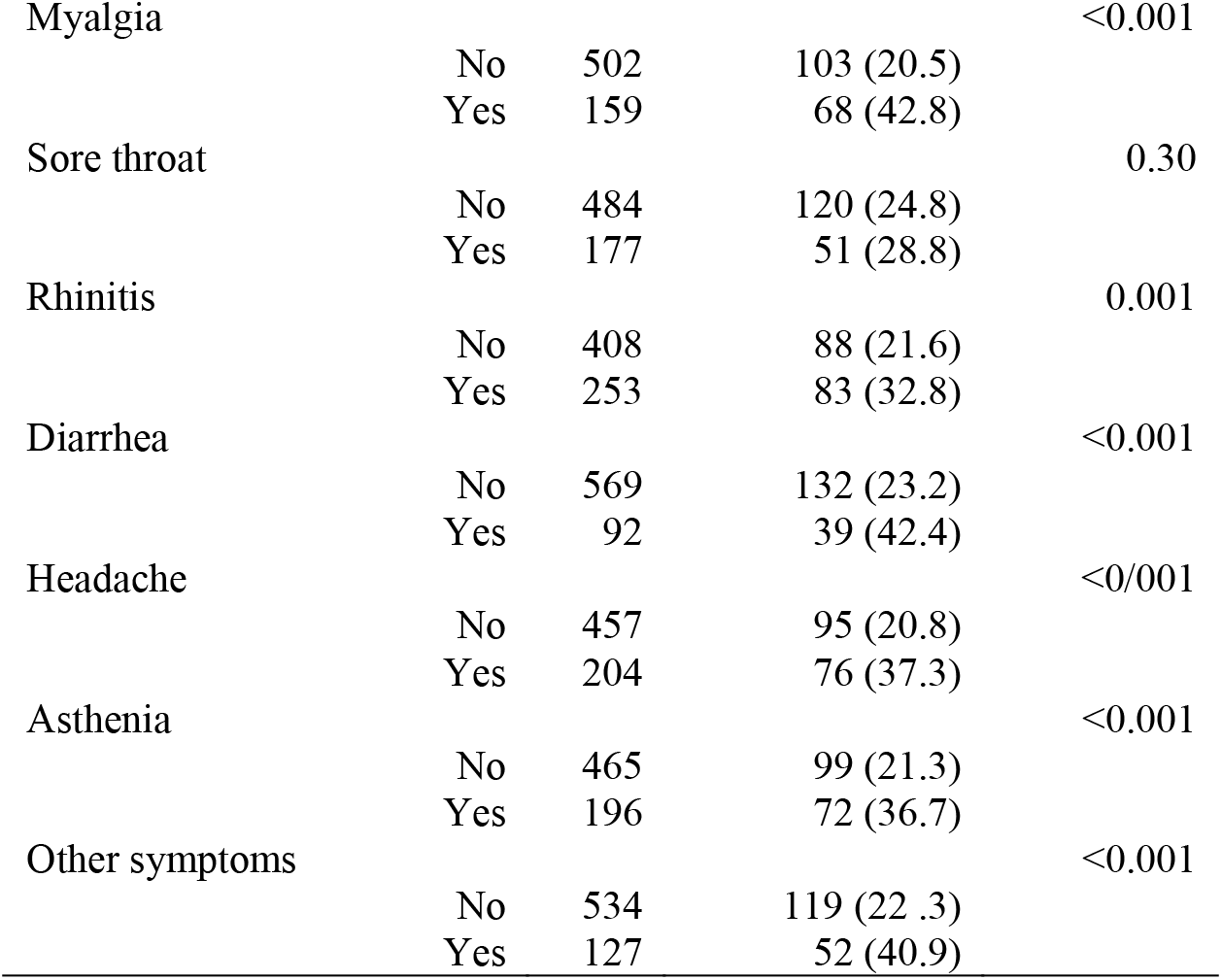
Proportion of participants with anti-SARS-CoV-2 antibodies

Participants who had experienced major symptoms were more likely to be infected, compared to those who had had minor or no symptoms (37.8%, 26.0%, and 13.9%, respectively, P <0.001). Of all symptoms considered, two had high positive predictive value for SARS-CoV-2 infection: anosmia (50/59 = 84.7%) and ageusia (52/59 = 88.1%). For all those who had anosmia and ageusia and negative serological findings, the time between reported symptoms and blood sampling was longer than one month, so the negative serological finding is unlikely to reflect delayed seroconversion.

Figure 2 displays the epidemic curve by week of onset of symptoms, serological status, and presence of major or minor symptoms. Among those with confirmed infection, the number of new cases dropped dramatically after week 7, corresponding to the beginning of the school holidays, and again after local confinement measures were introduced in the Oise department. As can be seen on Figure 2B, the symptoms among those who were not infected suggest that other respiratory viruses were circulating in that community during the study period.

**Figure 2.**
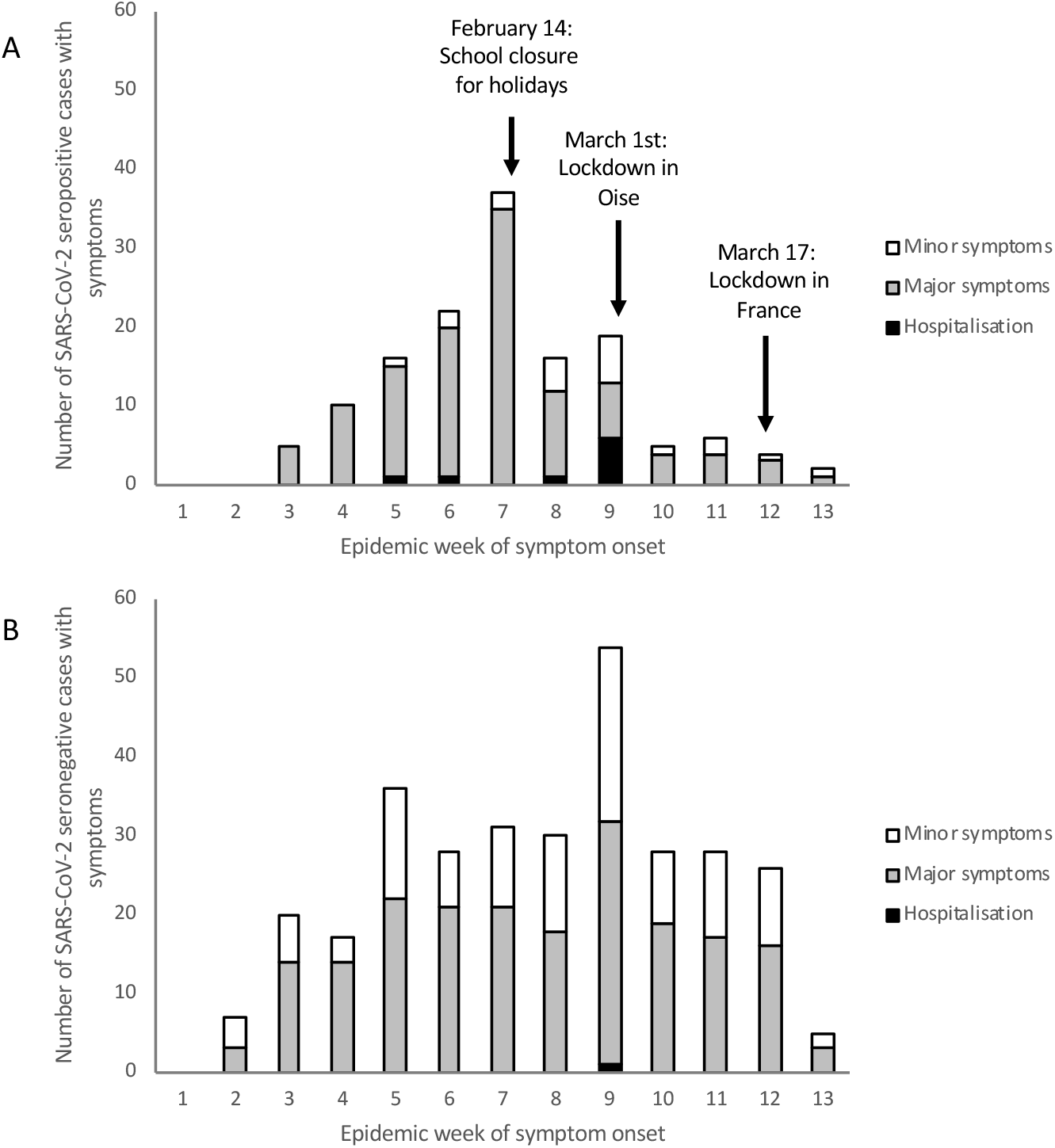
Timeline of symptom onset among (A) 142 symptomatic individuals who were seropositive for anti-SARS-CoV-2 antibodies and (B) 310 symptomatic individuals who were seronegative for anti-SARS-CoV-2 antibodies.

### Seroprevalence investigation among blood donors

Of the 200 serum samples that were collected from two blood donation centres between 23 and 27 March, 6 (3.0%, 95%CI = 1.1.- 6.4) had anti-SARS-CoV-2 antibodies.

## Discussion

This is, to our knowledge, the first study estimating by antibody detection the IAR of SARS-CoV-2 infection in a community affected by COVID-19, and the fist description of a COVID-19 outbreak in a school. In this cluster, we estimated that the IAR was 25.9%, approximately 8 weeks after the most likely introduction of SARS-CoV-2 circulation in this community. The epidemic presumably started during the third week of January, continued until school closure for holidays on February 15, and declined further following the introduction of confinement measures on March 1, with few confirmed cases on week 13.

The IAR was highest among the high school staff, teachers and pupils, and much lower among the parents and siblings of pupils. The secondary IAR in households was similar among parents (11.4%) and siblings (10.2%), and was close to the 15% observed in a study from Shenzen, China.^8^ In the context of the ongoing debates around the contribution of children and schools to viral transmission, these findings are of interest, and the impact of the school closure on the epidemic dynamic is particularly striking, with the limitation that these findings are restricted to high school setting only. High school pupils and their teachers had comparable IAR, similar to what was observed in Iceland when comparing the viral detection proportion between the 10-19 years old and adults in the targeted testing group.^9^

In the infected study population, the overall hospitalisation rate was 5.3%, and no death was observed. These relatively low figures need to be considered against the young age of a large part of the study population (40% were between 15 and 17 years of age; median age was 37 years; only two were older than 65 years). The current understanding of COVID-19 severity is that clinical presentation, hospitalisation rate and the CFR are lower among younger age groups.^3,10,11^

The IAR was higher among those with major symptoms, compared to those with minor or no symptoms. As documented before, anosmia and ageusia had high predictive values for COVID-19.^12,13^ While 17% of the infected had no symptoms during the study period, the true proportion of asymptomatic SARS-CoV-2 infections is likely higher since symptoms among those infected may also be attributable to the other respiratory viruses that were circulating in the community during that time period.

Smokers had a lower IAR compared to non-smokers. The association remained after adjustment for age or occupation. Earlier studies in China and the U.S. have documented a low proportion of smokers among COVID-19 patients (6% of 191 hospitalised patients in Wuhan^14^, and 1% of 7162 patients in the U.S.^15^). The protection associated with smoking in our study was very substantial (75% decrease in risk of infection), and deserves full attention. One possible explanation would be the downregulation of ACE2, the SARS-CoV-2 receptor^16^, by nicotine^17^. Such findings need replication, a solid understanding of the physiopathological process underlying it, and careful consideration in light of the increased risk of severe form of COVID-19 among smokers once infected^18^, and the long-term harmful consequences of smoking.

The main limitation of the study is the relatively low participation rate among all those invited. It is difficult to speculate whether this has led to an overestimation (if those who felt well during the study period did not come) or an underestimation (if those who had PCR-confirmed COVID-19 diagnosis during the study period did not come) of the IAR. Nevertheless, the overall findings were consistent with the literature to date in terms of the overall age-standardised CFR, hospitalisation rate, predictive values of symptoms such as ageusia and anosmia, and the possible effect of school closure on respiratory viruses epidemic dynamics.^3,9,10^ The clinical findings of our investigation were also limited by the fact that symptoms were retrospectively self-reported, in the absence of clinical evaluation, and that other respiratory viruses were presumably circulating at that time in the study population.

The choice of serological test has been a challenge in the absence of validated assays. We selected three serological methods with high (>99%) specificity, so that a positive signal with any of the three tests would be considered as a true positive.^6^ This, combined with the very high sensitivity of one of the three methods, the S-Flow assay, suggests that we were able to capture most if not all individuals with antibodies. A further concern relates to the antibody kinetics of SARS-CoV-2 infection. Most patients seroconvert within two weeks after onset of symptoms, but it is not clear whether time to seroconversion may be longer in patients with asymptomatic or subclinical infection.^5^ In such case, our estimate of the IAR might be an underestimate of the true one. Still, with a lockdown in place more than four weeks before the blood sampling, we believe that time to seroconversion has been sufficient for the majority of infected individuals. One important knowledge gap remains the extent to which the antibodies detected in this study would be immunoprotective. In the absence of this knowledge, we suggest that barrier measures and social distancing continue to apply the same way for people with and without antibodies.

We also found 3% of blood donors residing in the surrounding area to have evidence of anti-SARS-CoV-2 antibodies. While this proportion may seem low, despite circulation of the virus in the region, this fraction is likely to be lower than the true seroprevalence among adults in the region for a number of reasons. Blood donors tend to be more “health conscious”; they are not allowed to give blood if they have been ill in the month before; and tend to refrain from donating blood if a family member was recently ill. Nonetheless, tracking seropositivity in blood donors over time gives an indication of the trend in antibody dynamics in the general population, and for this reason, has been used effectively in previous epidemics, including pandemic H1N1 influenza^19^ and Zika virus.^20^ Repeated serologic testing for anti-SARS-COV-2 antibodies in blood donor populations needs to continue, in both affected and unaffected areas.

Overall, the findings of this study have important implications for public health measures and outbreak investigations specific to COVID-19. The overall IAR of 25.9% in a community living within the epicentre of the epidemic in France, and the low prevalence of antibodies among blood donors in the vicinity suggest that herd immunity will not be established quickly. Other areas of France, where the virus has not circulated, remain immunologically naive to the virus. Since 17 March 2020, the French government has implemented public health measures to restrict movement of individuals outside their home. The findings of this study suggest that lifting these confinement measures may be long and complex. For this reason, effective therapeutics treatments and vaccines specific to SARS-CoV-2 are urgently needed.

## Data Availability

The authors confirm that the data supporting the findings of this study are available within the article [and/or] its supplementary material.

## Contributors

AF, BH, SC, and PG designed the investigation.

IC, CD, LK managed the collection of data on-site.

NJ, SFP, MNU oversaw the adherence of the study to the regulatory requirements.

LT, CB oversaw the collection of the data and maintained the database.

ME, OS, SVDW, CD, LG, ST and TB developed the serological testing.

ST, KYC, BC, SM, IS, LG performed the serological analyses

LT, YM performed the statistical analyses.

RG, AF and BH drafted the first versions of the manuscript.

All authors critically reviewed and approved the final version of the manuscript.

## Acknowledgements

We would like to thank the Directorate General for Health for facilitating the initiation of the study, Santé publique France for linking with their investigation team, the Agence Régionale de Santé des Hauts de France and the Academia of Amiens for their continuous support throughout the realization of the study. We would also like to thank Nathalie De Parseval, Claire Dugast, Valentine Garaud, Soazic Gardais, Caroline Jannet, Fanny Monboisse, Isabelle Porteret, Hantaniaina Rafanoson, Sandrine Ropars, Nicole Corre-Catelin, Laurence Arowas, and Mamou Traore who participated in the organisation and the implementation of the field investigation. Finally, we would like to thank Fabrice Carrat and Dominique Costagliola for critical reading of the manuscript and for their useful comments.

## Supplementary Material

Major symptoms included fever, cough, dyspnea, anosmia and ageusia; minor symptoms included sore throat, rhinitis, myalgia, diarrhea, headache and asthenia.

**Supplementary Material Table 1.** Seropositivity according to symptom severity and serological assay performed on serum samples from 661 participants

|  | Total<br>(n=661) | No symptoms<br>(n=209) | Minor only<br>(n=131) | Major<br>(n=321) |
| --- | --- | --- | --- | --- |
| Binding | 164 (24.8) | 27 (12.9) | 19 (14.5) | 118 (36.8) |
| ELISA | 96 (14.5) | 13 (6.2) | 10 (7.6) | 73 (22.7) |
| LIPS | 136 (20.6)* | 20 (9.6)* | 16 (12.2) | 100 (31.1) |
| Any positive | 171 (25.9) | 29 (13.9) | 21 (16.0) | 121 (37.7) |
| Combination of the<br>3 serologies |  |  |  |  |
| None positive | 490 (74.1) | 180 (86.1) | 110 (84.0) | 200 (62.3) |
| 1 positive | 35 (5.3) | 11 (5.3) | 6 (4.6) | 18 (5.6) |
| 2 positives | 47 (7.1) | 5 (2.4) | 6 (4.6) | 36 (11.2) |
| 3 positives | 89 (13.5) | 13 (6.2) | 9 (6.9) | 67 (20.9) |

\*1 sample with no result

